# Characterizing Adulterant and Polysubstance Use Research Priorities through Syringe Residue Analysis in Kentucky

**DOI:** 10.64898/2026.07.17.26358092

**Authors:** Kathleen R. McNealy, Preston Tolbert, Mike Ward, Kelli Harpe, Kacey Byczek, Cassandra D. Gipson, Amanda Fallin-Bennett, Rachel A. Vickers

## Abstract

Polysubstance use is rising and linked to heightened overdose rates and increased treatment challenges, further exacerbated by increasing detection of adulterants (e.g., xylazine) in the street drug supply. Harm reduction groups provide sterile syringes in exchange for used ones, creating a unique opportunity to characterize prevalent polysubstance combinations and inform translational and preclinical research We analyzed residues from used syringes (N=3,168) obtained from several harm reduction organizations in Jefferson County, KY (Jan-Dec 2025) for the presence of substances using gas chromatography mass spectrometry (GC-MS). We classified compounds as adulterants (e.g., diphenhydramine [DPH]/Benadryl), byproducts/precursors of synthesis (e.g., 4-ANPP), and recreational drugs (e.g., meth). We excluded byproducts/precursors and determined the most frequent substance and pairs/trios containing one or more recreational substance. Results. Of 3,168 syringes, 2,522 (79.61%) tested positive for substances. Out of those positive, the top recreational substances were meth (n=1,387; 54.99%), fentanyl (n=1,220; 48.37%), and heroin (n=653; 25.89%). Top adulterants were DPH (n=1021; 40.48%), dimethyl sulfone (n=749; 29.69%), and lidocaine (n=736; 29.18%). The most common pairs were DPH+fentanyl (n=670; 26.57%), lidocaine+fentanyl (n=659; 26.13%), dimethyl sulfone+meth (n=621; 24.62%), and fentanyl+heroin (n=484; 19.19%). The most common trios were DPH+lidocaine+fentanyl (n=369; 14.63%), DPH+fentanyl+heroin (n=327; 12.97%), lidocaine+fentanyl+heroin (n=297; 11.77%), diphenhydramine+xylazine+fentanyl (n=273; 10.82%), and meth+lidocaine+fentanyl (n=262; 10.39%). Our findings highlight evolving patterns of multiple-opioid and opioid-stimulant polysubstance use, generating insights that can be rapidly applied to strengthen clinical, preclinical, and translational polysubstance research. These insights allow for investigations into biobehavioral mechanisms and consequences of emerging use patterns, accelerating development of novel therapeutics.

## Introduction

United States opioid overdose rates remain high and methamphetamine-involved overdose deaths are rising^1^. Further, over 30% of overdose deaths now involve opioid-stimulant co-use^1^. These crises now intersect within an increasingly complex street drug supply, with the growing presence of psychoactive adulterants (e.g., xylazine) that can increase overdose rates, withdrawal, and adverse health outcomes^2^. Thus, there is a critical need to determine what substances co-occur in the street drug supply to guide researchers in characterizing drivers and harms of polysubstance use and adulteration of the street drug supply.

Drug checking programs, such as point-of-care sample testing or syringe residue analysis, provide opportunities for researchers to learn about real-world use patterns. Existing syringe residue reports in the scientific literature are often focused on describing prevalence of individual substances and adulterants, or specific, a priori substance pairs^3,4^, limiting their utility in informing polysubstance research. Here, we analyze syringe residues from an urban Kentucky county to identify prevalent polysubstance combinations.

## Methods

Briefly, from January-December 2025, residue from 3,168 used syringes collected in partnership with five community-based harm reduction organizations in Jefferson County, Kentucky were qualitatively analyzed via gas chromatography-mass spectrometry (GC-MS). We classified resulting compounds as primary agents defined as recreationally used substances, *adulterants* defined as psychoactive compounds not typically associated with recreational use, *bulking agents* defined as compounds not typically associated with psychoactive effects but added to increase product volume, or contaminants defined as precursors/byproducts of illicit drug synthesis (see Table 1 and Supplementary). Metabolites were grouped with their associated primary agent. To focus on epidemiologically relevant patterns, we excluded *contaminants* and identified the most prevalent substance pairs and trios involving the top three observed primary substances. Notably, these pairs and trios reflect substances commonly co-occurring, but syringes may contain additional compounds. We also report the frequency that each of the top substances occurred in the absence of other non-contaminant substances.

**Table 1.** Top 10 primary agents, adulterants, and bulking agents.

| **Substance** | **N**<br>**Substance-**<br>**Positive** | **%**<br>**Substance-**<br>**Positive** | **N Mono-**<br>**Substance** | **% Mono-**<br>**Substance** |
| --- | --- | --- | --- | --- |
| ***Any Substance*** | 2522 | 79.61 | 469 | 18.59 |
| ***Primary Agents*** |  |  |  |  |
| Methamphetamine | 1387 | 43.78 | 330 | 23.79 |
| Fentanyl | 1220 | 38.51 | 87 | 7.13 |
| Heroin* | 622 | 19.63 | 5 | 0.80 |
| Tramadol | 213 | 6.72 | 2 | 0.94 |
| Cocaine* | 188 | 5.93 | 8 | 4.26 |
| Gabapentin | 175 | 5.52 | 7 | 4.00 |
| Morphine | 137 | 4.32 | 4 | 2.92 |
| Codeine | 96 | 3.03 | 0 | 0.00 |
| Amphetamine | 77 | 2.43 | 0 | 0.00 |
| Parafluorofentanyl | 34 | 1.07 | 0 | 0.00 |
| ***Adulterants*** |  |  |  |  |
| Diphenhydramine | 1021 | 32.23 | 33 | 3.23 |
| Lidocaine | 736 | 23.23 | 12 | 1.63 |
| Xylazine | 464 | 14.65 | 11 | 2.37 |
| Medetomidine | 364 | 11.49 | 1 | 0.27 |
| Caffeine | 224 | 7.07 | 2 | 0.89 |
| Acetaminophen | 172 | 5.43 | 1 | 0.58 |
| Dextromethorphan | 46 | 1.45 | 0 | 0.00 |
| Carbinoxamine | 17 | 0.54 | 0 | 0.00 |
| Quinine | 14 | 0.44 | 0 | 0.00 |
| Levamisole | 10 | 0.32 | 0 | 0.00 |
| ***Bulking Agents*** |  |  |  |  |
| Dimethyl Sulfone | 749 | 23.64 | 11 | 1.47 |
| D-mannitol | 400 | 12.63 | 0 | 0.00 |
| Glycerin | 29 | 0.92 | 0 | 0.00 |
| Sorbitol | 4 | 0.13 | 0 | 0.00 |
| Cholesterol | 2 | 0.06 | 0 | 0.00 |
| Sesamin | 2 | 0.06 | 0 | 0.00 |

*Note:* *N syringes with substance* reflects the number of syringes testing positive for the specified substance. *% syringes with substance* is calculated using all syringes testing positive for any substance as the denominator. *N mono-substance syringe* reflects the number of syringes containing only the specified substance. *% mono-substance* is calculated using all syringes testing positive for that substance as the denominator. \*indicates that those syringes contained either the drug and/or drug metabolites

## Results

See Table 1 for overall and mono-substance single compound frequencies in the tested syringes. Of 3,168 syringes tested, 79.61% syringes contained at least one substance. Only 18.59% of substance-containing syringes had a single non-contaminant substance, with the remaining containing two (21.17%), three (14.55%), or up to 18 non-contaminant substances. Top substances in each category are reported in Table 1. Methamphetamine, fentanyl, and heroin were the most common primary drugs, occurring in 43.78%, 38.51%, and 19.63% of syringes, respectively. The most prevalent substance combinations involving each and how often each appeared alone are visualized in Figure 1. Methamphetamine was the sole substance in 23.79% of methamphetamine-positive syringes. Fentanyl and heroin were detected alone far less frequently (7.13% and 0.80%, respectively). Methamphetamine most commonly co-occurred with dimethyl sulfone, diphenhydramine (DPH), fentanyl, and lidocaine respectively. Fentanyl most frequently co-occurred with 1) DPH, 2) lidocaine, 3) heroin, 4) xylazine, and 5) methamphetamine, while heroin most commonly occurred with 1) fentanyl, 2) DPH, 3) lidocaine, and 4) medetomidine.

**Figure 1.**
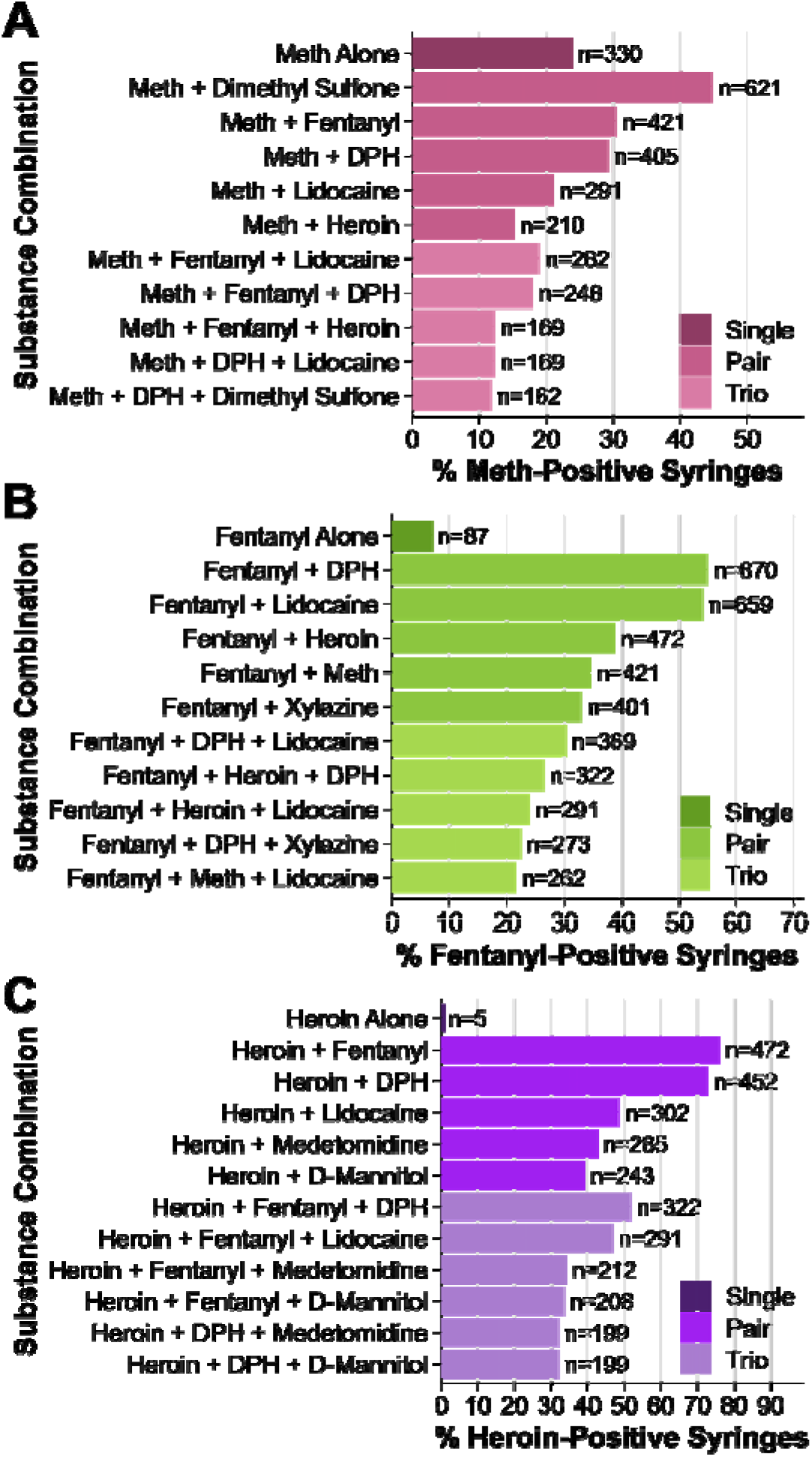
Frequency of substance-alone and co-occurring substances among top primary agent-positive syringes. Horizontal bar plots show the percentage of syringes testing positive for methamphetamine (A), fentanyl (B), or heroin (C) that also contained additional substances. Each panel displays the prevalence of the primary agent alone and the top five pairs and trios involving that substance. Substance combinations are ordered by prevalence and combination size within each panel. Values on bars indicate the number of positive syringes for each substance combination. Percentages are calculated using the total number of syringes positive for the primary substance in each panel as the denominator (see Table 1). The syringe may contain additional compounds beyond the two⍰ or three⍰substance combinations shown, and therefore the total percentages exceed 100%.

## Discussion

Methamphetamine occurred alone most frequently, consistent with reports of less adulterated methamphetamine versus opioid supplies^5^. Nonetheless, all primary substances occurred with other substances more than they appeared alone. Our observed high rates of fentanyl detection and an impure opioid supply^3^, continued opioid adulteration with veterinary anesthetics such as xylazine or medetomidine^2^, and opioid-methamphetamine co-occurrence^1^ align with national trends.

Antihistamine DPH (Benadryl) and local anesthetic lidocaine were amongst the most prevalent adulterants with methamphetamine, fentanyl, and heroin. DPH has long been noted as an opioid adulterant^6^ and is associated with heightened overdose rates^7^. However, DPH with methamphetamine is not frequently reported (but see^4^) and concerning given preclinical reports of antihistamines increasing methamphetamine potency^8^. Notably, a follow-up analysis indicated that 28.9% of methamphetamine + DPH syringes contained no opioid, supporting that DPH co-occurrence with methamphetamine was not solely attributable to concurrent opioid presence. Similarly, lidocaine has been widely detected with cocaine^9^, but not in opioids or methamphetamine as in the present report. Notably, lidocaine and DPH are used clinically to lower the therapeutic dose of opioids by enhancing opioid potency^10^, and thus likely exert similar enhancing effects of street opioids, warrenting further study. The extent to which three or more substances appeared in syringes also underscores the need to understand unique harms resulting from complex mixtures. For example, given high prevalence of fentanyl with DPH *and* lidocaine and their like opioid-sparing effects, the two simultaneously interacting with fentanyl could pose significant health harms. When discussing our results with a local community advisory board of people with living experience using drugs, adulterant modulation of substance dependence, withdrawal, and overdose emerged as important polysubstance-related outcomes for future study.

Importantly, we cannot determine whether a single syringe reflects one drug supply or rather multiple drug supplies resulting from repeated use of the same syringe. These data also reflect a single, urban county in Kentucky and are pooled across a full year. Thus, although many of our findings were consistent with longstanding national trends, our results may not generalize to other areas or timepoints. Nonetheless, our findings provide new insights into prevalent co-occurring substance combinations while presenting a novel approach for reporting syringe residue data to inform polysubstance research.

## Supporting information

Supplemental Methods for Research Letter

## Data Availability

De-identified data produced in the present study are available upon reasonable request to the authors.

## Acknowledgements and Funding Sources

We would like to thank the Survivors Union of the Bluegrass for their careful input on preliminary results and future research priorities. This publication was supported by the National Center for Research Resources and the National Center for Advancing Translational Sciences, National Institutes of Health, through Grant UL1TR001998 (UK Center for Clinical and Translational Sciences [CCTS]). Other funding sources include: R01 DA058933 (CDG); UK CCTS SUD Pilot (RAV, AFB, & CDG); Vital Strategies (RAV & AFB). KRM was fully supported by TL1TR001997 while preparing this manuscript). The content is solely the responsibility of the authors and does not necessarily represent the official views of the NIH nor the University of Kentucky.

## Notes

### Competing Interest Statement

The authors have declared no competing interest.

