## Supplemental Methods for Research Letter for "Characterizing Adulterant and Polysubstance Use Research Priorities through Syringe Residue Analysis in Kentucky"

### **Supplementary Methods**

#### **Sampled Material**

Syringes were collected in partnership with five community-based harm reduction organizations in Jefferson County, KY. Collected syringes were delivered to the lab in dedicated sharps bins and labeled with the site, zip code (if applicable), date delivered to lab/date collected from the site, and a unique letter identifier if multiple bins were collected from the same site on the same day (ex: A, B, C, etc.). Upon delivery to the lab, the sharps container was opened and the syringes tipped onto a flat surface where 1-2 batches of up to 48 syringes per site (maximum batch size for GC-MS) were then selected randomly for analysis using laboratory tongs/forceps. Syringes were only excluded for the following reasons: 1.) if syringes had a broken barrel, 2.) if syringes lacked a syringe plunger, or 3.) if syringes contained a large amount of blood. The first two criteria were implemented to prevent contamination of substances from other syringes, and the latter to prevent damage to the gas chromatography/mass spectrometer (GCMS).

#### **Laboratory Analysis**

##### **Sample Preparation**

Selected syringes were lined up on a piece of lab paper. The plungers were removed from the syringes, and 0.5 mL of methanol was poured into each syringe barrel. The plungers were then reinserted and used to expel the methanol into clean, labeled autosampler vials containing 0.5 mL of methanol with proadifen (SKF-525A; 10 ng/mL) as an internal standard. For syringes that were clogged and could not be emptied by depressing the plunger, the plunger was removed and the methanol was poured from the rear of the syringe barrel into the corresponding autosampler vial and used for residue analysis in the GC-MS. Following solvent transfer, the syringes and plungers were discarded in an approved sharps waste container.

### **Analytical Methods**

Residues are then analyzed via GCMS. This method allows a wide range of substances to be detected (including novel substances); all the substances we detected are listed below in the Substance Classification section. Controls were utilized to determine validation parameters (limit of detection, reproducibility, accuracy, specificity, etc.). GCMS analysis was conducted using a 30-meter HP-5MS UI column with an Agilent 8860 Gas Chromatograph and an Agilent 5977 MSD mass spectrometer and the National Institute of Standards and Technology (NIST) spectral library. We used a temperature program from 80 °C (hold 1 minute) to 260 °C (hold 10 minutes) at a ramp rate of 20 °C per minute.

**Substance Classification**

After samples were analyzed, we listed the detected compounds and noted whether the compound was present or absent in each syringe (qualitative analysis). Over the course of the data collection timeline, we identified 84 total substances that we classified into primary/recreationally used agents, adulterants, bulking agents, and contaminants according to commonly used classification methods^1,2^ according to the definitions below.

Primary/Recreationally Used Agents: Drugs with pharmacological activity that are sought out and injected to attain specific effects. Note that some compounds classified here may also function as adulterants. However, because the intended purpose of their inclusion cannot be determined from qualitative toxicological data alone, classification is based on their known pharmacological activity and their potential to be intentionally used as drugs.

- Methamphetamine
- Fentanyl
- Heroin & Diacetylmorphine (Heroin)
- Tramadol
- Cocaine
- Gabapentin
- Codeine
- Amphetamine
- Para Fluorofentanyl
- Ketamine
- Carfentanil
- Dronabinol
- Methocarbamol
- Clonazepam
- Valeryl Fentanyl
- Mephedrone
- Hydrocodone
- Ortho Fluorofentanyl
- Metonitazene
- Methadone
- Nordazepam
- Pentobarbital
- Buprenorphine
- Morphine

Adulterants/Psychoactive Adulterants: Compounds displaying pharmacological effects that may alter the effects of the primary drug.

- Diphenhydramine (Antihistamine)
- Lidocaine (Local anesthetic)
- Xylazine (Veterinary anesthetic)
- Medetomidine (Veterinary anesthetic)
- Caffeine (Stimulant)
- Acetaminophen (NSAID)
- Carbinoxamine (Antihistamine)
- Levamisole (Immunomodulating agent/Anti-parasitic)
- Doxylamine (Antihistamine)
- Dexmedetomidine (Veterinary anesthetic)
- Ibuprofen (NSAID)
- Dextromethorphan (Anti-tussive)
- Guaifenesin (Expectorant)
- Noscapine (Anti-tussive)
- Promethazine (Antihistamine)
- Benzyl alcohol (Local anesthetic)
- Naloxone (Overdose reversal)
- Tetracaine (Local anesthetic)
- Quinine (Antimalarial)
- Epiquinine (Antimalarial)
- Estradiol Valerate (Hormone)
- Progesterone (Hormone)
- Melatonin (Hormone)
- Benzocaine (Local anesthetic)
- Papaverine (Muscle relaxant)
- Btmps

Bulking Agents: Compounds not typically associated with psychoactive effects added to increase the yield or quantity of the drug at the point of sale.

- D-mannitol (Artificial sweetener)
- Sorbitol (Artificial sweetener)
- Dimethyl sulfone
- Glycerin
- Cholesterol
- Sesamin

Contaminants: These are components that occur as a result of illicit synthesis and are described as manufacturing byproducts.

- X4_Anpp
- X4_Piperidinol_2_2_6_6_Tetramethyl
- Acetylcodeine
- Diethylene_Glycol_Dibenzoate
- N_Butyl_2_2_6_6_Tetramethyl_4_Piperidinamine
- Acetanilide_N_1_Phenethyl_4_Piperidyl
- Benzyl_Chloride
- X2_5_Dimethoxy_4_Chloro_Fentanyl
- Despropionyl_Ortho_Methylfentanyl_2
- N_Propionyl_Norfentanyl
- Ecgonine
- X2_5_Dimethoxy_4_Ethylthio_Fentanyl
- Tropacocaine
- Phenethyl_4_Piperidone
- Despropionyl_Ortho_Methylfentanyl
- X1_Phenethyl_4_Piperidone_2
- X2_5_Dimethoxy_3_4_Dimethyl_Fentanyl
- X2_5_Dimethoxy_4_Ethyl_Fentanyl
- Oxazepam_2tms_Derivative_Quality_55
- Emdp_Quality_64
- X2_5_Dimethoxy_Fentanyl
- X2_4_Dimethoxy_Fentanyl
- X2_5_Dimethoxy_4_Isopropylthio_Fentanyl
- X2_Methyl_Fentanyl
- X2_5_Dimethoxy_Fentanyl_2
- Despropionyl_Para_Fluorofentanyl

Metabolites: Compounds that result from drug metabolism/breakdown, resulting from instability of the drug or the individual retracting blood into the syringe.

- Norcocaine (Cocaine)
- Monoacetylmorphine (Heroin)
- Methyl Ecgonine (Cocaine)
- Benzoyl Ecgonine (Cocaine)
